# Optimal blood glucose targets for critically ill patients with sepsis in the intensive care unit

**DOI:** 10.1101/2025.11.03.25339362

**Authors:** Gousia Habib, Kevin Soon Hwee Teo, Willem van den Boom, Mengling Feng, Kay Choong See

**Affiliations:** Saw See Hock School of Public Health (SSHSPH), National University of Singapore; Division of Neurology, Department of Medicine, National University Hospital, Singapore; Institute for Human Development and Potential, Agency for Science, Technology and Research, Singapore; Division of Respiratory and Critical Care Medicine, Department of Medicine, National University Hospital, Singapore

**Keywords:** Sepsis, ICU, Blood Glucose Management, Glycemic Control, Mortality Risk

## Abstract

**Background:** Hyperglycemia and hypoglycemia are key risk factors for morbidity and mortality in critically ill septic patients. Despite ongoing research, glucose management guidelines for critically ill patients remain inconsistent, especially for diabetic patients. This study identifies optimal glucose targets to reduce mortality in critically ill patients with sepsis.

**Research Question:** What is the optimal blood glucose range associated with reduced in-hospital mortality for critically ill patients with sepsis, and how does this vary by diabetic status?

**Study Design and Methods:** This cohort study analyzed 22,374 adult intensive care unit (ICU) patients with sepsis from the MIMIC-IV database. Non-linear logistic regression models assessed the relationship between 72-hour median blood glucose levels and in-hospital mortality, adjusting for age, gender, and Sequential Organ Failure Assessment (SOFA) score. Subgroup analyses explored variations based on diabetic status and other clinical factors.

**Results:** The study found a U-shaped relationship between blood glucose levels and mortality, with the lowest risk at 6.3 mmol/L overall and 6.8 mmol/L for diabetic patients. A 5–8 mmol/L glucose range during the first 72 hours was associated with a mortality risk below 10%, representing up to a 5% reduction in mortality compared to the guideline targets of 7.8–10.0 mmol/L. No significant differences were found between patients with or without skin and soft tissue infection.

**Interpretation:** The findings suggest that a tighter glycemic control range of 5–8 mmol/L could improve survival in ICU patients with sepsis, challenging current guidelines. Further randomized controlled trials are necessary to validate and optimize glycemic control strategies for critically ill septic patients.

**Clinical Trial Registration:** Not applicable.

**Take-Home Points:** *Study Question:* What is the optimal blood glucose level for reducing mortality in critically ill patients with sepsis?

*Results:* A U-shaped relationship was found, with the lowest mortality risk at 6.3 mmol/L overall and 6.8 mmol/L for people with diabetes. Patients with a 5–8 mmol/L glycemic range had an approximate 50% reduced mortality risk compared to those who maintained glycemic ranges within existing guideline recommendations.

*Interpretation:* Unlike the guideline-recommended glucose targets of 7.8 - 10mmol/L, a tighter glycemic target (5–8 mmol/L) may improve mortality for critically ill patients with sepsis.

## Introduction

Glycemic control in critically ill patients is a topic of clinical interest, due to the prevalence of diabetes mellitus in the adult population and recognition that hyperglycemia and hypoglycemia are common but deleterious metabolic derangements in critically ill patients with sepsis [1]. Hyperglycemia may alter leukocyte-endothelial interactions and increase the production of proinflammatory factors and reactive oxygen species, which result in direct cellular damage. In contrast, hypoglycemia indicates depletion of glycogen reserves and failure of gluconeogenesis to meet peripheral tissue needs [2, 3]. Increased glycemic variability may also be associated with increased mortality in critically ill patients and is thought to be the result of increased oxidative stress, endothelial dysfunction and apoptosis [4].

The optimal glucose target for critically ill patients in the intensive care unit remains under debate, with a relative lack of consensus between published guidelines. For example, a consensus guideline 2009 by the American Diabetes Association and American Association of Clinical Endocrinology recommended a glucose target between 7.8– 10mmol/L for the general population of patients in intensive care [5]. A more recent guideline by the Society of Critical Care Medicine, published in 2024, recommended against a lower glucose target of 4.4–7.7 mmol/L over a glucose target of 7.8–11.1 mmol/L for critically ill adults [6]. These recommendations follow the NICE-SUGAR trial that reported increased morbidity and mortality in critically ill patients who underwent stringent glycemic control through intensive insulin therapy [6,7].

While growing evidence indicates that blood glucose is positively associated with mortality in critically ill non-diabetic patients, patients with diabetes subjected to tight glucose control may be exposed to greater risk due to relative hypoglycemia [8]. However, the LUCID trial found no significant difference in mortality for diabetic patients assigned to a liberal glucose target of 10.0–14.0 mmol/L versus a target range of 6.0–10.0 mmol/L, contradicting guidelines recommending 7.8–10.0 mmol/L for the general population of critically ill patients [9, 10].

Given the ongoing uncertainty, we sought to analyse the relationship between glucose levels and mortality for intensive care unit (ICU) patients with sepsis. The study aimed to define an optimal glucose range for critically ill diabetic and non-diabetic patients, as well as specific subgroups of interest which have clinical associations with chronic hyperglycemia, such as patients with chronic liver disease, chronic kidney disease, heart failure, cancer, and soft-tissue skin infections.

## Materials and Methods

### Data Description

The Medical Information Mart for Intensive Care (MIMIC)-IV database version 3.1 [11,12,13] served as our data source. The database contains 73,181 ICU admissions of adults above 18 at Boston’s Beth Israel Deaconess Medical Centre from 2008 to 2019. Managed by the Laboratory for Computational Physiology at the Massachusetts Institute of Technology, MIMIC-IV contains hourly physiological measurements from bedside monitors, patient demographics, diagnoses using ICD-9/10 codes, and additional clinical information gathered during standard medical care. The database features comprehensive documentation and publicly available code from its user community. It adheres to the ethical guidelines of the Massachusetts Institute of Technology’s institutional review board (no. 0403000206) and the 1964 Helsinki declaration with subsequent amendments.

We used the following inclusion criteria to subset the MIMIC-IV database: patients were included based on sepsis within a 72-hour window (24 hours before and 48 hours after ICU admission) from the Sepsis-3 derived table of MIMIC-IV [6]. The Sepsis-3 criteria identified sepsis based on suspected infection and concurrent organ dysfunction (e.g., SOFA score ≥2) [14]. The exclusion criteria were patients who were readmitted to the ICU or did not have blood glucose measurements in the first 72 hours of the ICU stay. Only the first ICU stay with sepsis of each patient was considered to ensure the analysis focused on the initial sepsis event.

Figure 1 illustrates the selection process for ICU stays. From an initial 94,458 ICU admissions, patients without sepsis and repeat admissions were excluded, resulting in 22,506 ICU admissions with sepsis. The characteristics of the patient cohorts identified through this selection process are presented in Table 1.

**Figure 1:**
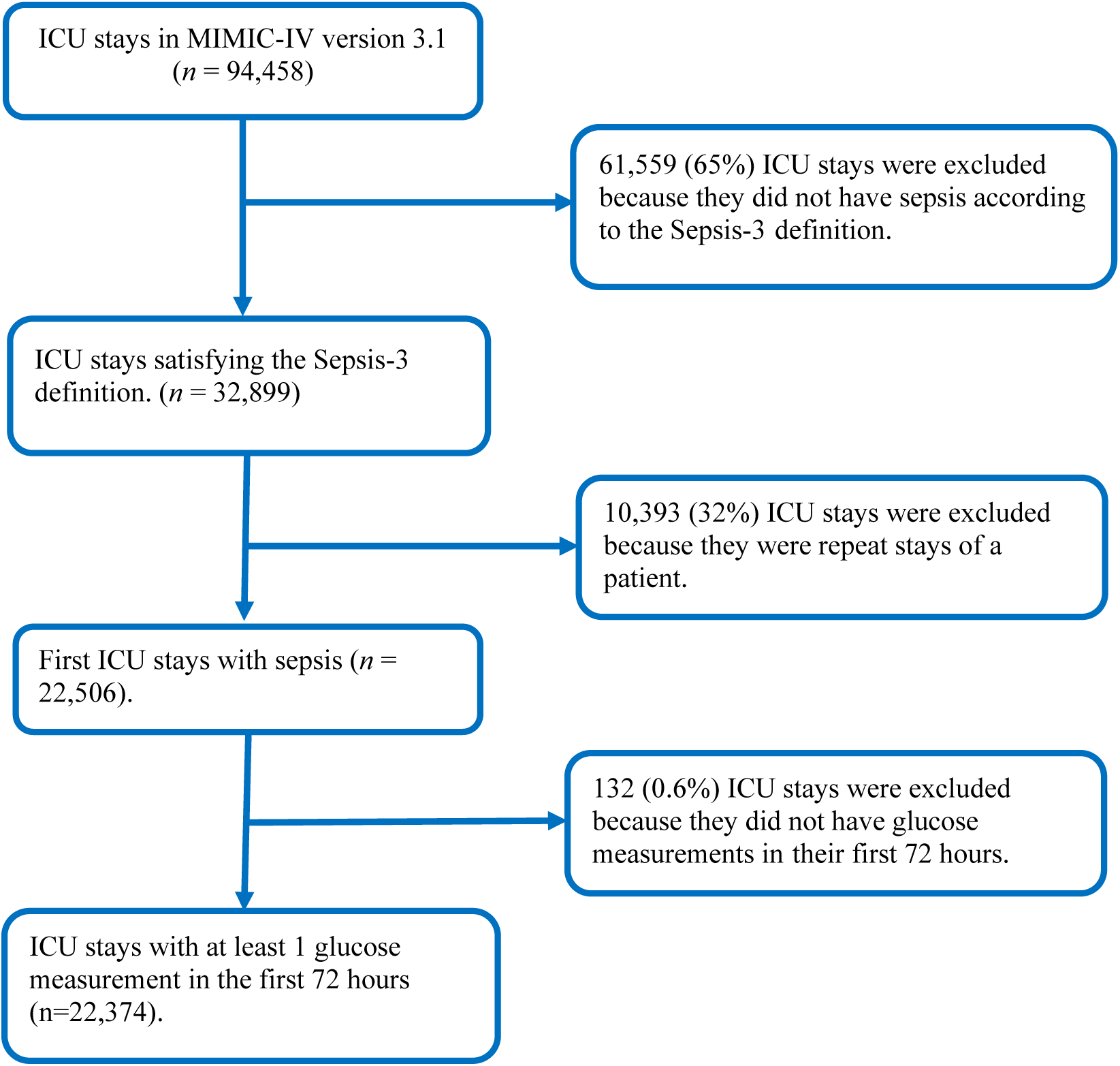
Case inclusion flowchart.

**Table 1:** Patient cohort characteristics.

| *Characteristic* | *Mean or count (%)* | *Quartiles* |  |  |
| --- | --- | --- | --- | --- |
|  |  | *Q1* | *Median* | *Q3* |
| Age (years) | 66.0 | 56 | 68 | 79 |
| Male gender | 12,953 (58%) |  |  |  |
| SOFA score* | 5.84 | 3 | 5 | 8 |
| In-hospital mortality | 3,416 (15%) |  |  |  |
| Median of the blood glucose measurements in the first 72 hours (mmol/L) | 7.5 | 6.0 | 7.0 | 8.2 |
| Number of glucose measurements in the first 72 hours | 7.0 | 4.0 | 6.0 | 9.0 |

\*SOFA: Sequential Organ Failure Assessment

### Data collection

Data collected from included subjects comprised demographic and clinical characteristics such as age, sex, Sequential Organ Failure Assessment (SOFA) score, diabetic status, comorbidities (chronic kidney disease (CKD), chronic liver disease (CLD), heart failure, cancer), vasopressor use, and renal replacement therapy. Glucose measurements were taken from blood gas and chemistry datasets in MIMIC-IV [11,12].

### Statistical Analysis

To assess the relationship between glycemia control approaches (tight glycemic control versus standard glycemic control and mortality in ICU patients with sepsis, we utilised a generalized additive model (GAM) [15]. Tight glycemic control and blood glucose levels are maintained within a stricter target range, with therapy adjusted more aggressively to achieve this goal. In contrast, standard glycemic control targets a broader blood glucose range, with fewer adjustments to treatment. This method was selected for its ability to model non-linear relationships to examine the effects of glucose levels on patient outcomes [16]—the model adjusted for age, gender and Sequential Organ Failure Assessment (SOFA) score. Using a GAM enabled us to identify clinically relevant thresholds for optimal glucose levels in septic patients, offering valuable insights into how glycemic control strategies influence patient outcomes.

We considered demographic factors, skin and soft tissue infections (SSTI), comorbidities, and clinical interventions for subgroup analyses. Specifically, we defined subgroups based on (i) age; (ii) gender; (iii) race; (iv) SSTI; (v) diabetes; (vi) cancer; (vii) chronic liver disease; (viii) heart failure; (ix) chronic kidney disease; (x) dialysis (or continuous renal replacement therapy); (xi) vasopressor use (norepinephrine, epinephrine or dopamine)..

Source code for the data extraction and statistical analyses can be found at https://github.com/gousiya26-I/Optimal-Glucose-Control-in-Sepsis [Will be made available upon publication].

## Results

This section presents the study cohort characteristics and the relationship between 72-hour median blood glucose levels and mortality risk in ICU sepsis patients. Subgroup analyses, including diabetic status and comorbidities such as chronic liver and kidney disease, are also discussed. The study cohort characteristics are described in Table 1.

### Median 72-hour blood glucose and mortality risk

In the undifferentiated study cohort, mortality risk exhibited a U-shaped relationship with median 72-hour blood glucose, with the lowest risk at a median glucose of 6.3 mmol/L (Figure 2). On subgroup analysis, the optimal median 72-hour glucose for diabetic patients was higher at 6.81 mmol/L and non-diabetic 6.19 mmol/L (e-Figure 4).

**Figure 2:**
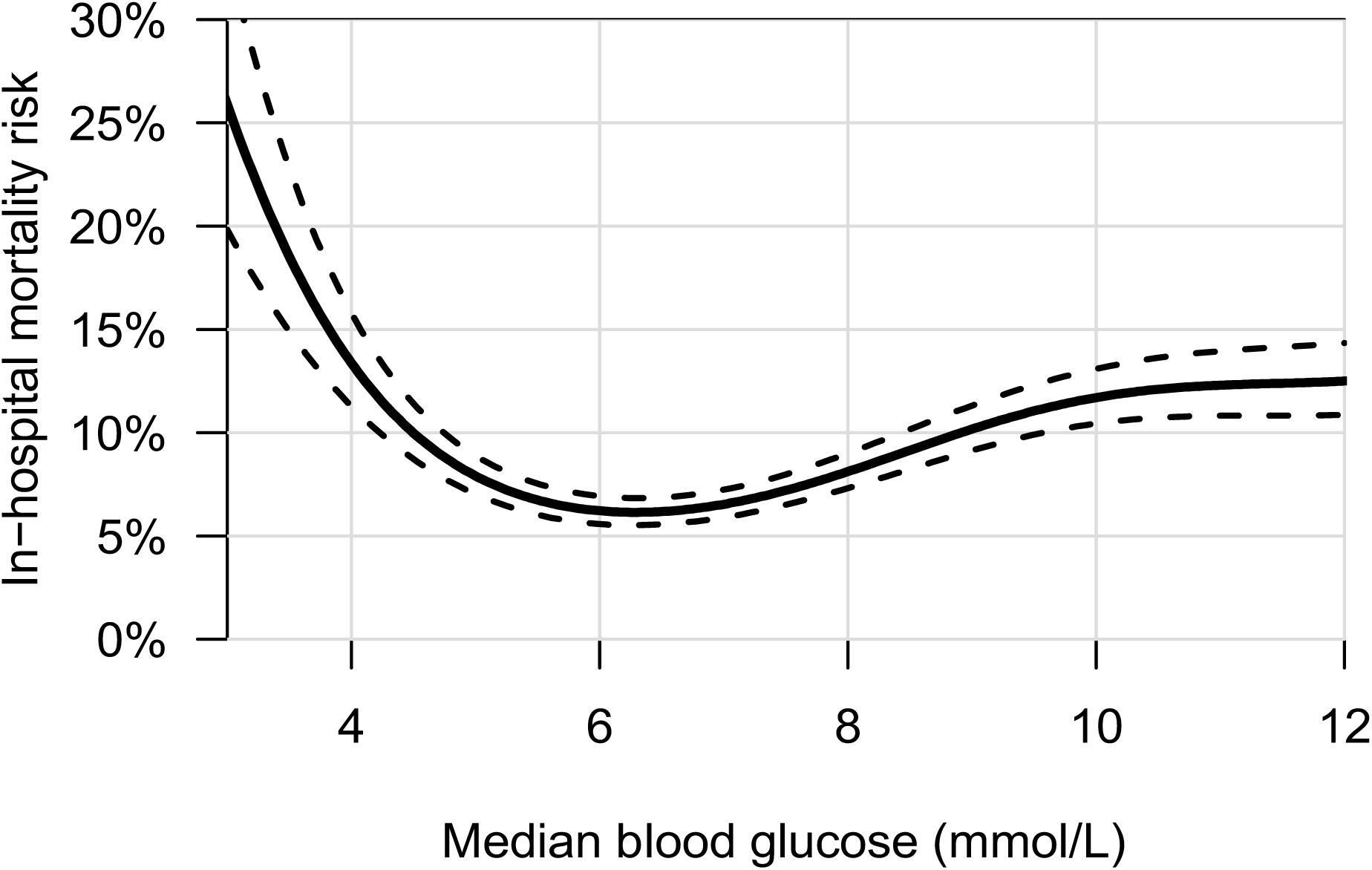
Association of 72-hour median blood glucose levels and in-hospital mortality risk for patients admitted to the ICU with sepsis. The solid line represents the estimated risk, and the dashed lines indicate the 95% confidence intervals.

**Figure 3:**
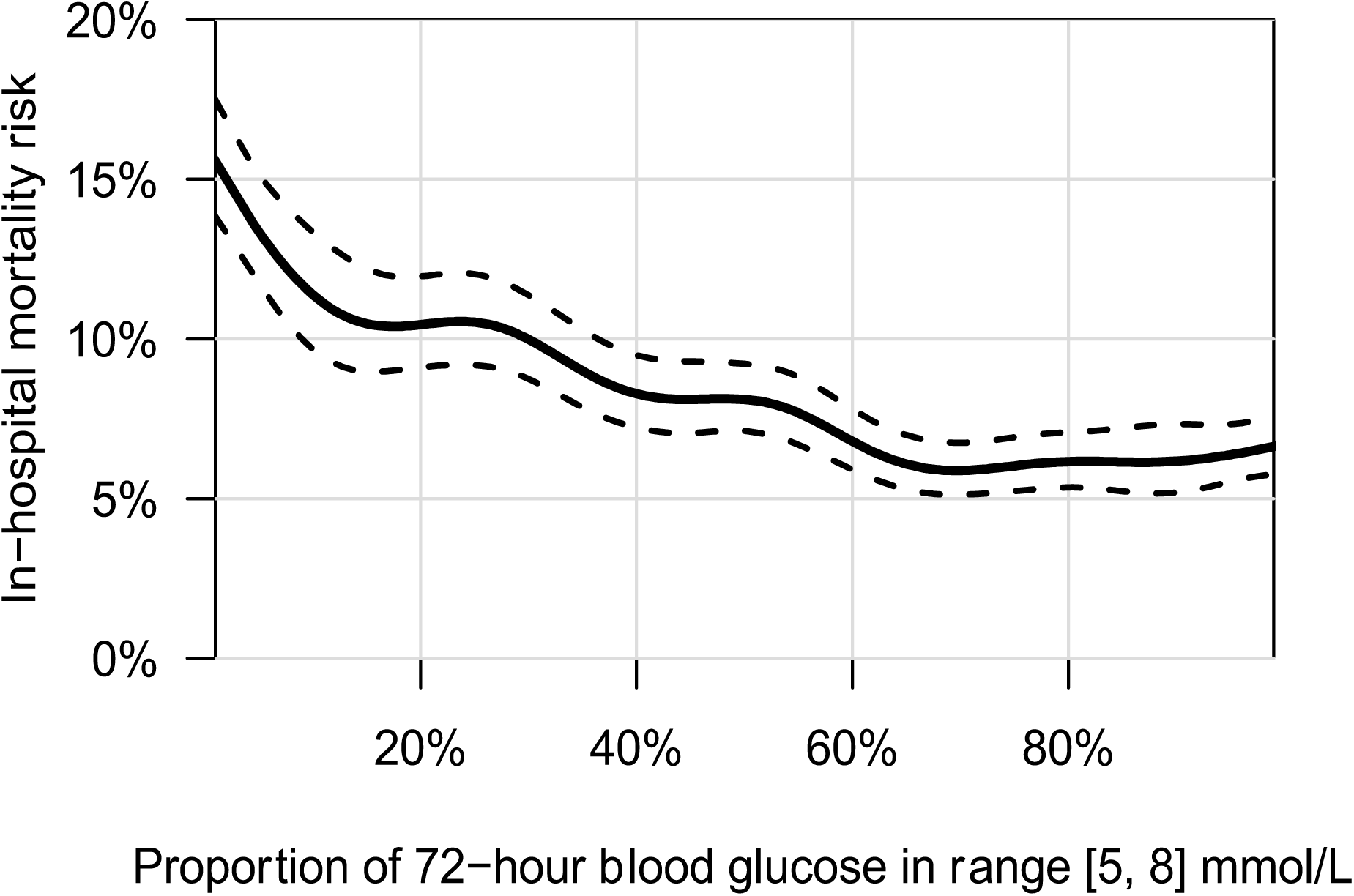
Association of the proportion of glucose measurements in the range [5, 8] mmol/L and in-hospital mortality risk. The solid line represents the estimated risk, and the dashed lines indicate the 95% confidence intervals.

**Figure 4:**
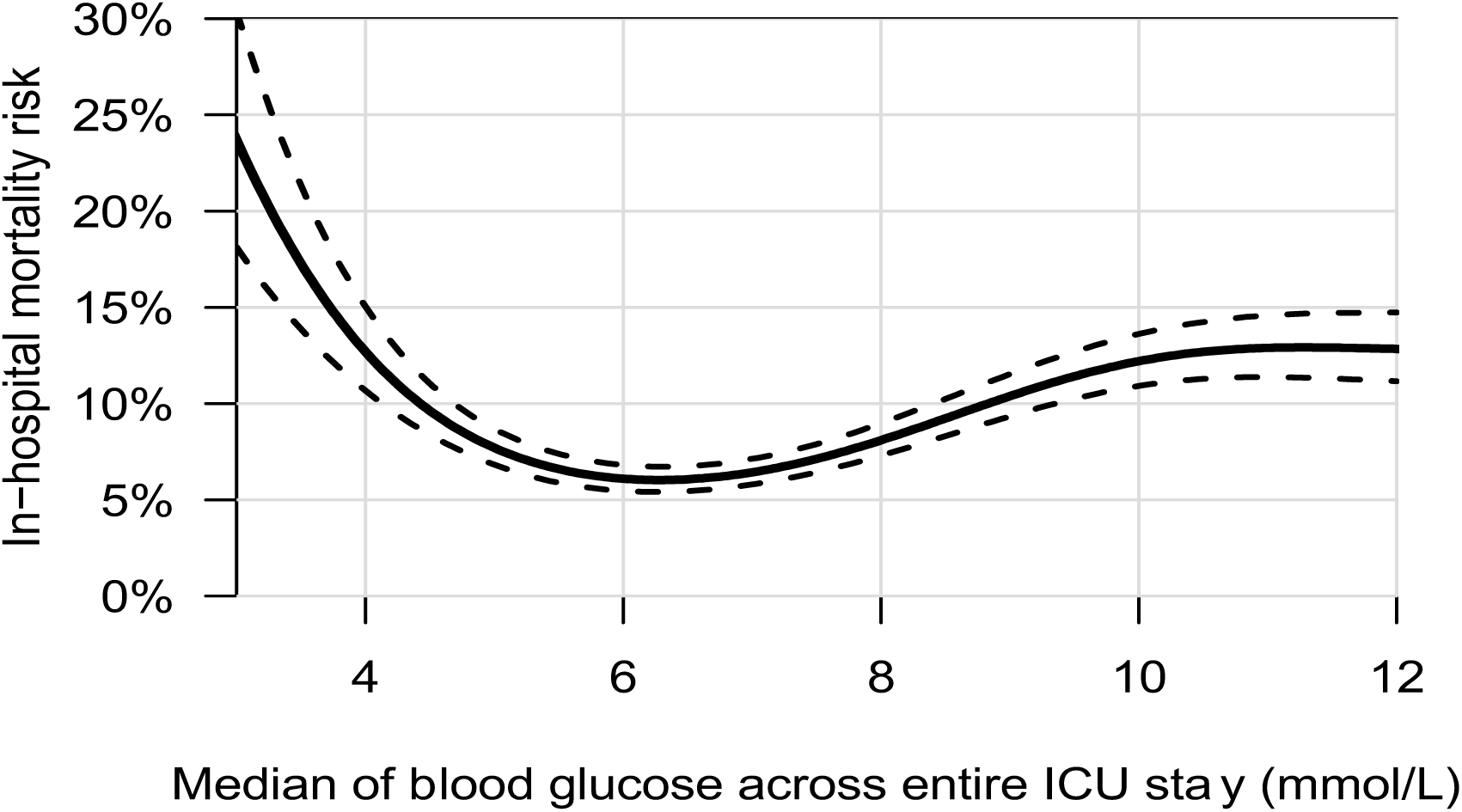
Association of median blood glucose levels over the entire ICU stay and in-hospital mortality risk. The solid line represents the estimated risk, and the dashed lines indicate the 95% confidence intervals.

The results of the subgroup analyses are presented in the supplementary material (e-Figure 1 to e-Figures 12). The optimal 72-hour glucose levels associated with the lowest mortality risk vary across subgroups. For instance, in patients with chronic kidney disease (CKD), the optimal glucose level is 6.64 mmol/L, and 6.61 mmol/L in those with heart failure (HF). However, it should be noted that specific subgroups contain too few ICU stays for accurate estimation, and we did not perform statistical tests for subgroup differences.

As a follow-up to Figure 2, we present another analysis on the relationship between blood glucose regulation and mortality risk in ICU patients. To account for clinical reality where fluctuations in glucose inevitably occur, we defined a 72-hour median glucose range of 5–8 mmol/L where the mortality risk remained below 10%. Specifically, we considered the percentage of glucose measurements that fall within the range of 5–8 mmol/L as a predictor in the GAM model for mortality risk, instead of the median glucose. Figure 3 shows that maintaining 80% of glucose measurements within this range, compared to 20%, is associated with a roughly 50% reduction in mortality risk (*p* < 0.001).

### Sensitivity Analysis

We conducted two sensitivity analyses. Firstly, we considered all glucose measurements, instead of only those from the first 72 hours, when computing the median glucose (Figure 4). Secondly, we used the mean instead of the median of the glucose measurements from the first 72 hours (Figure 5). The U-shapes in Figures 4 and 5 are consistent with those in Figure 2, considering the median glucose of the first 72 hours.

**Figure 5:**
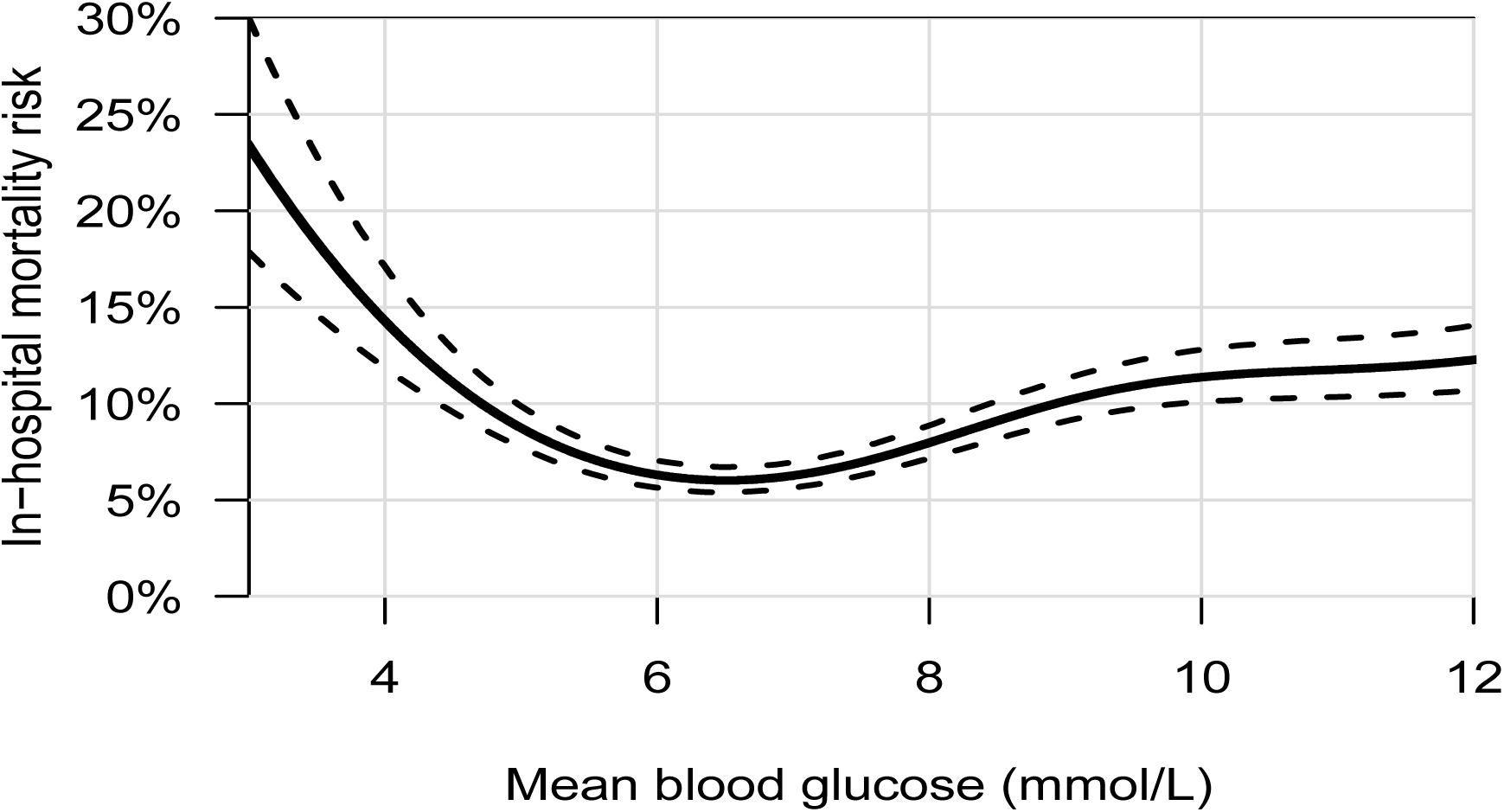
Association of 72-hour mean blood glucose levels and in-hospital mortality risk. The solid line represents the estimated risk, and the dashed lines indicate the 95% confidence intervals.

## Discussion

Using a large real-world dataset from the MIMIC-IV database, our study investigated the association between blood glucose levels and mortality in critically ill patients with sepsis admitted to the ICU. The results revealed a U-shaped relationship between 72-hour median blood glucose and in-hospital mortality, with the lowest risk observed at 6.3 mmol/L. Notably, an increasing mortality risk is seen in patients whose 72-hour median blood glucose was below 5.0 mmol/L, corroborating previous studies which have identified a greater burden of mortality risk in patients with sepsis and hypoglycemia [17]. The association between increased mortality and low blood glucose may be due to hypoglycemia being a biomarker of disease processes that confer a predisposition to adverse outcomes, including liver and kidney failure, adrenal failure and malnutrition. Hypoglycemia may also directly increase mortality by mediating the release of inflammatory molecules and impairing autonomic homeostasis and white cell responses [18]. While less marked than hypoglycemia, the deleterious effect of hyperglycemia is also evident by a near-doubling of the mortality risk for patients with a median blood glucose of 10 – 12 mmol/L as opposed to 6.3 mmol/L. Hyperglycemia alters leukocyte-endothelial interactions, resulting in direct cellular damage through increased production of proinflammatory factors and reactive oxygen species [2].

The optimal median 72-hour glucose for diabetic patients was higher at 6.8 mmol/L with a flattened curve gradient. The difference in optimal glucose targets for existing diabetic patients may indicate adaptations to chronic hyperglycemia and explain why patients with pre-existing diabetes did not benefit from tight glucose control in previous randomized controlled trials [17, 19, 20].

We identified an optimal glucose range of 5–8 mmol/L during the first 72 hours of ICU admission, within which mortality risk remained below 10%. Patients who maintained glucose within the optimal range (5–8 mmol/L) for at least 80% of the first 72 hours of their ICU stay had a significantly reduced mortality rate compared to those who maintained glucose within this range for less than 20%. This demonstrates that consistent glucose control within this range may improve survival outcomes in septic ICU patients. Our subgroup analyses indicated that the optimal glucose target may vary based on diabetes status. We did not find convincing evidence for a notable difference across other subgroups. The lack of significant variation suggests the glucose target range could be broadly applicable, although individualization based on comorbidities and clinical conditions remains critical.

Specific subgroups’ distinct metabolic challenges and pathophysiological mechanisms are reflected in their higher optimal glucose levels, such as 6.64 mmol/L in those with chronic kidney disease (CKD).. In CKD patients, renal dysfunction impairs the kidneys’ ability to filter and excrete excess glucose, contributing to increased insulin resistance and glucose intolerance. This necessitates higher glucose targets to prevent hypoglycemia and maintain metabolic stability [23]. Note that for specific subgroups, especially those with limited sample sizes (e.g., CLD, dialysis patients and certain racial/ethnic groups), the broader 95% confidence intervals indicate increased uncertainty regarding the estimations of ideal glucose levels. Smaller datasets introduce increased variability, resulting in less accurate estimations. The key strengths of the current study include the use of a large real-world dataset from the MIMIC-IV database, which enhances the generalizability of the findings to a vast population of critically ill septic patients. Additionally, identifying the U-shaped relationship between blood glucose and mortality provides valuable insights for improving glycemic control strategies in intensive care.

However, this study has some limitations. As an observational study, there is a potential for residual confounding from unmeasured variables that may affect the results. Observational studies cannot establish direct cause-and-effect relationships, and retrospective data may introduce biases. Additionally, the glucose measurements used in the study were not continuous, which may impact the accuracy of the findings. Although the cohort is large, individual patient variability and the use of insulin or other medications were not fully accounted for in our analysis.

These findings require further validation, and future research could explore machine learning and deep learning techniques to better predict patient outcomes based on glucose levels and other clinical data. Moreover, generalizability of the findings should be tested by replicating the study in databases from different demographic regions, such as Europe, China, and Korea. Integrating continuous glucose monitoring (CGM) and closed-loop insulin systems could enhance glycemic control in critically ill patients. Furthermore, future randomized controlled trials could compare a glucose range of 5-8 mmol/L against conventional glycemic targets of 7.8–10.0 mmol/L in critically ill patients with sepsis, ensuring strict adherence to glycemic ranges using real-time intra-arterial glucose monitoring and closed-loop systems.

## Conclusion

This study reveals a U-shaped relationship between blood glucose levels and mortality in critically ill patients with sepsis, whereby the lowest mortality risk was associated with a glucose target of 6.3 mmol/L and a glycemic range of between 5–8 mmol/L. This glucose range is lower than many current guidelines, particularly for non-diabetic ICU patients, highlighting the need for reassessing current glycemic control protocols and more individualized glucose management in the ICU setting. Further randomized controlled trials are necessary to validate and optimize glycemic control strategies for critically ill septic patients.

## Acknowledgements

The authors would like to thank Keiko Asao, Xi Zhi Low and Krittaphas Chaisutyakorn for their assistance with the data analysis.

## Author contributions

MF and KCS conceived the study. Gousia and WvdB curated the data. Gousia and WvdB performed the data analysis. Gousia, Kevin and WvdB produced an initial draft of the manuscript. All authors provided feedback on the manuscript. Gousia and WvdB had full access to all the data. All authors had final responsibility for the decision to submit for publication.

## Additional information

The e-Figure and e-Tables are available under the Supplementary Material document provided separately.

## Abbreviations

ICU: Intensive Care Unit
SOFA: Sequential Organ Failure Assessment
SSTI: Skin and Soft Tissue Infection
MIMIC-IV: Medical Information Mart for Intensive Care
IV ICD: International Classification of Diseases
GAM: Generalised Additive Model
RCT: Randomised Controlled Trial CGM Continuous Glucose Monitoring

## Supplementary Information to

### 1. Subgroup analyses

This section considers subgroup analyses based on demographic factors, skin and soft tissue infections (SSTI), comorbidities, and clinical interventions. We aim to determine how these variables affect the association between blood glucose levels and in-hospital mortality. Specifically, we consider subgroups based on (i) age; (ii) gender; (iii) race; (iv) SSTI; (v) diabetes; (vi) cancer; (vii) chronic liver disease; (viii) heart failure; (ix) chronic kidney disease; (x) dialysis (or continuous renal replacement therapy); (xi) vasopressor use (norepinephrine, epinephrine or dopamine). Vasopressors are considered since they are frequently utilised in critically ill patients to sustain blood pressure, and their application may influence the mortality risk related to blood glucose levels.

Septic patients with potential skin-related infections, i.e. SSTI, were identified by ICD codes starting with ‘68’ [1]. The comorbidities were identified using the following ICD-9/ICD-10 codes:

Diabetes: 250, E10, E11 and T21 [2]

Cancer: C, D0, D1, D3, D4 [3]

Chronic liver disease: 571, K70, K73, K74, K76 [4]

Heart Failure: 428, I50 [5]

Chronic kidney disease: 585, N18 [6]

The subgroup analyses use the 72-hour median blood glucose as in the main analysis. e-Figures 1 to 12 contain the results. Largely, there are no notable difference between subgroups, where it should be noted that certain subgroups have a small number of cases resulting in high statistical uncertainty.

---

**e-Figure 1:**
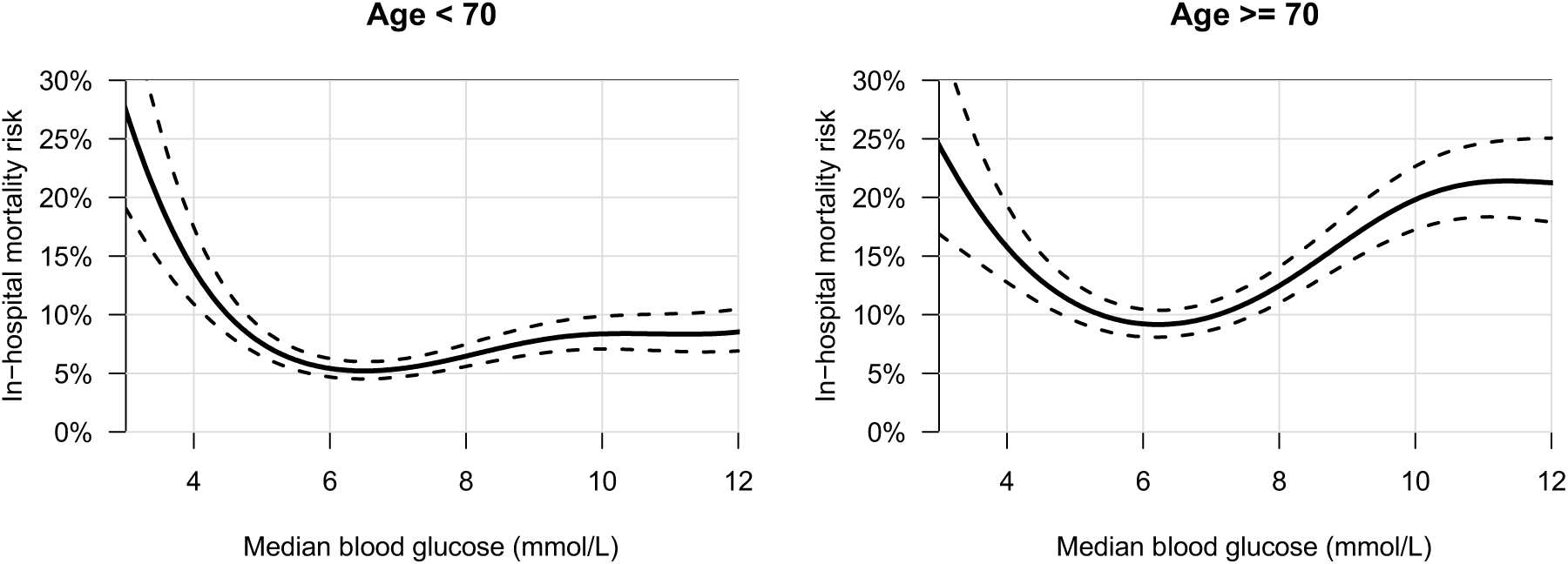
Subgroup analysis based on age. < 70 years old (n = 12,239) versus ≥ 70 years old (n = 10,135)

**e-Figure 2:**
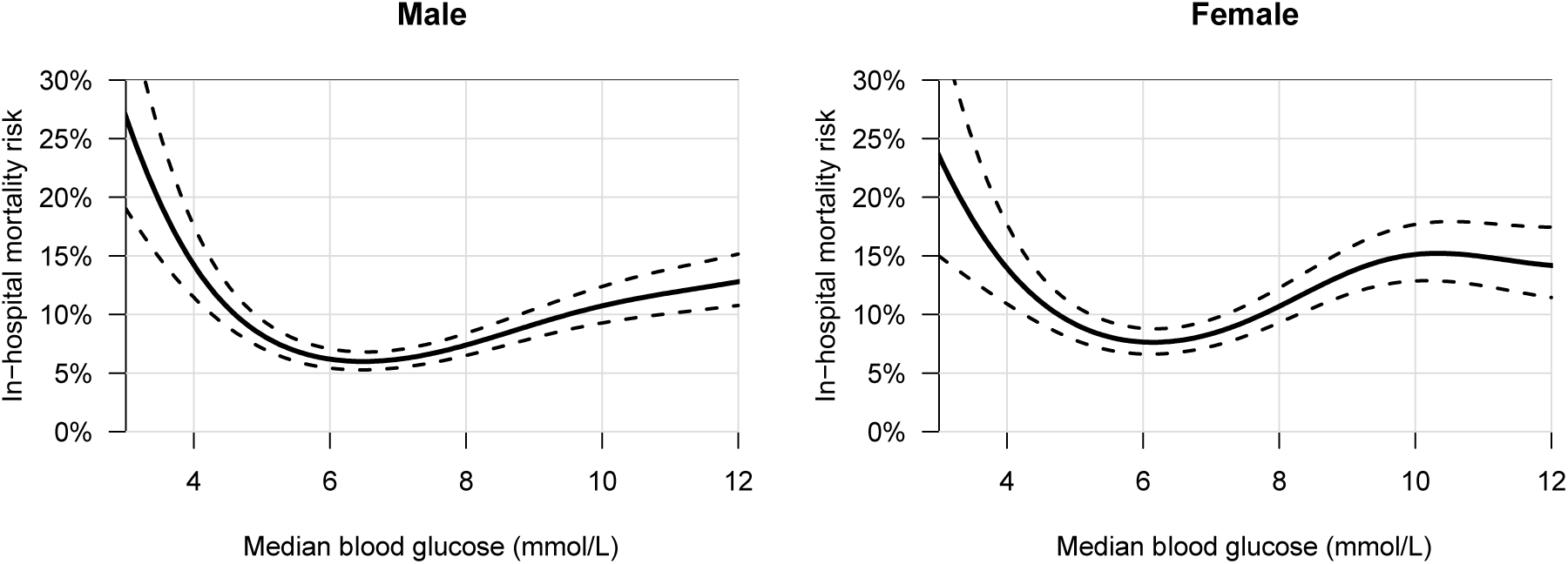
Subgroup analysis based on gender. Male (*n* = 12,953) versus female (*n* = 9,421)

**e-Figure 3:**
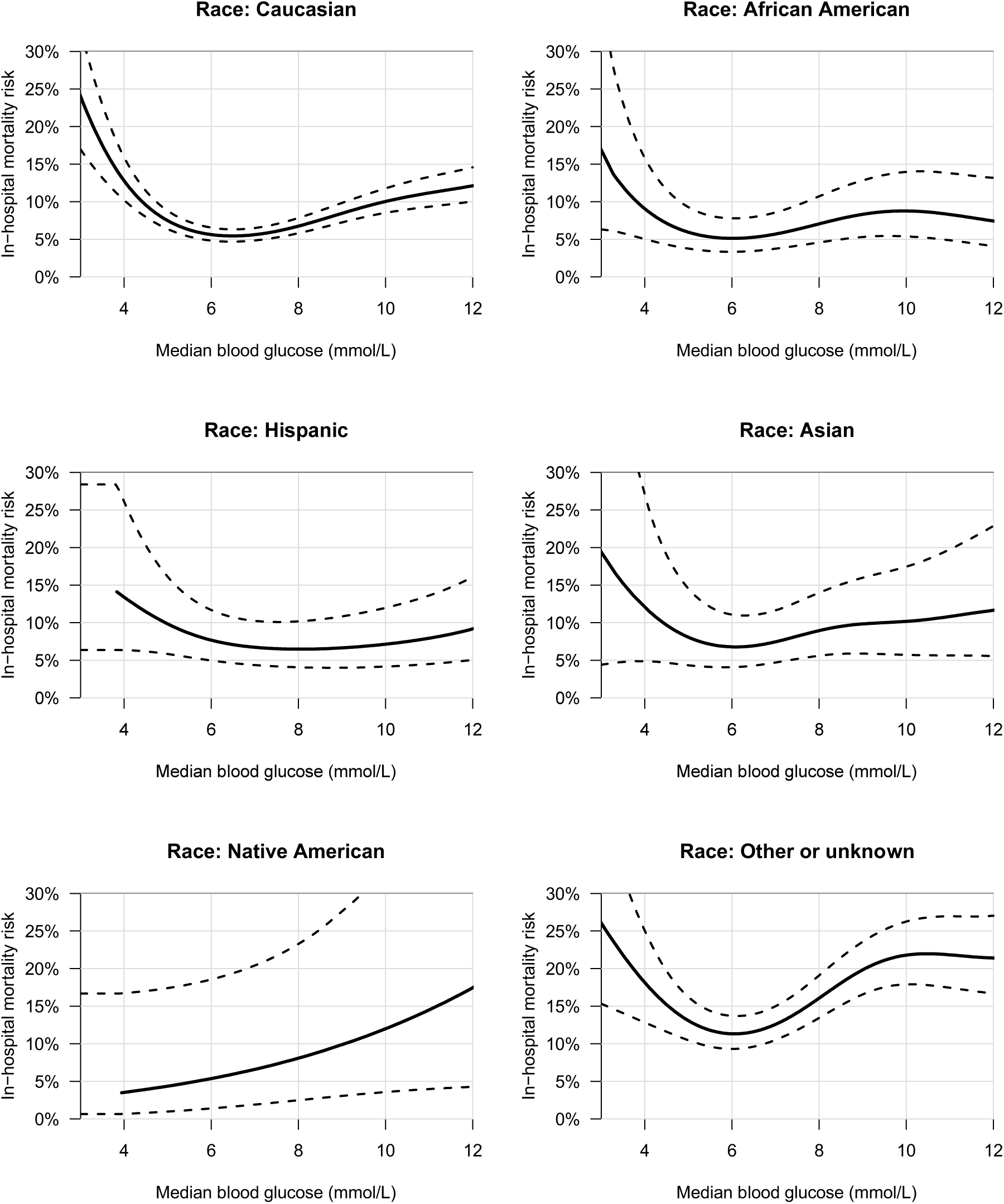
Subgroup analysis based on race Caucasian (*n* = 15,076), African American (*n* = 1,511), Hispanic (*n* = 722), Asian (*n* = 645), Native American (*n* = 82), Other or unknown (*n* = 4,338)

**e-Figure 4:**
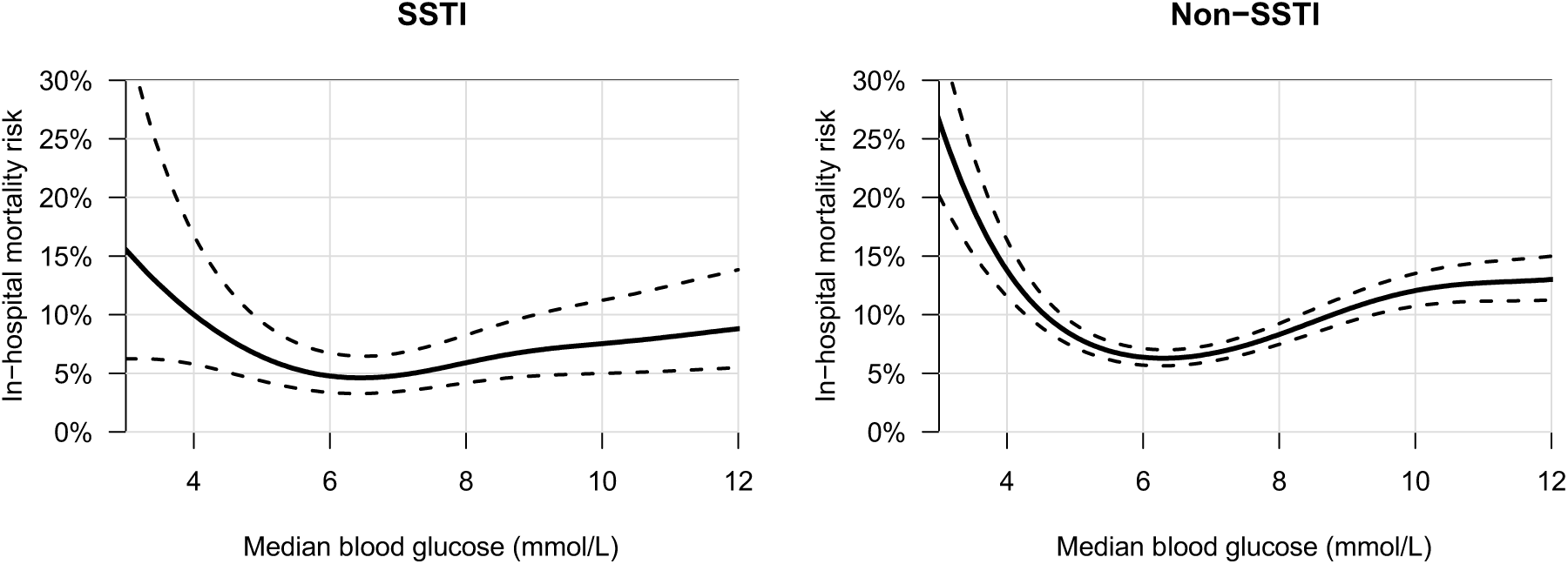
Subgroup analysis based on skin and soft tissue infections (SSTI) SSTI (*n* = 1,655) versus non-SSTI (*n* = 20,719)

**e-Figure 5:**
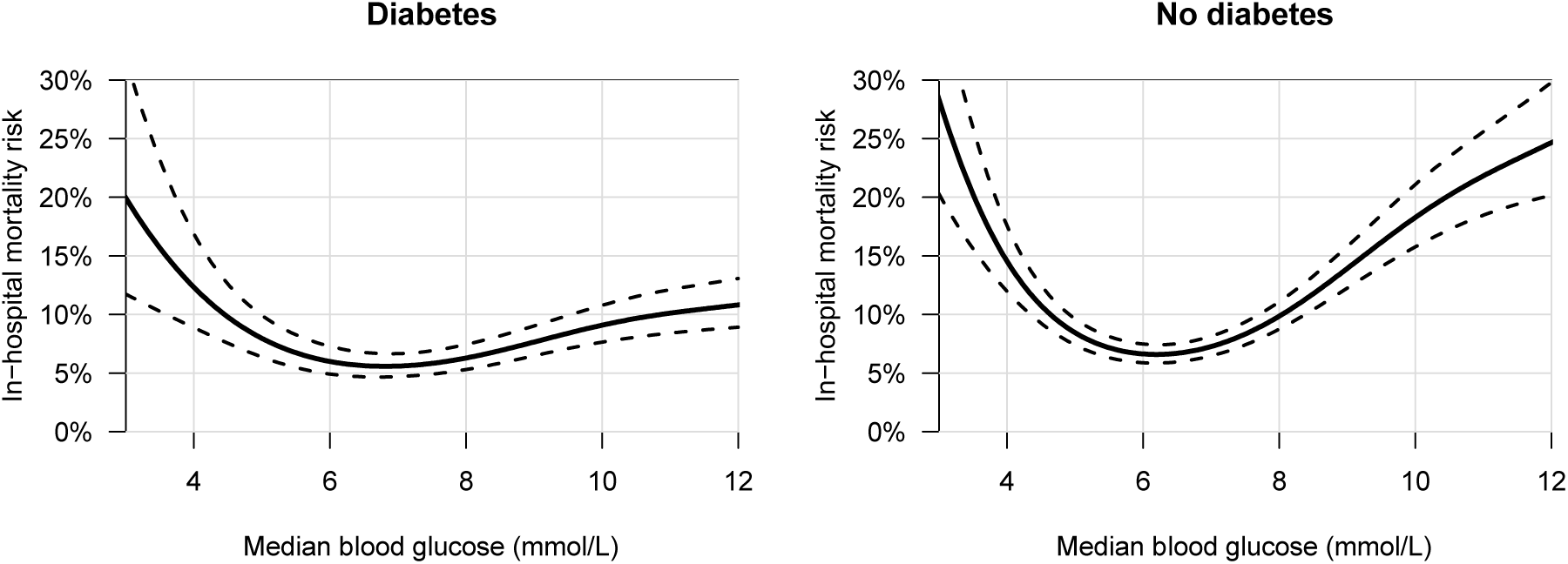
Subgroup analysis based on diabetes. Diabetes (n = 6,701) versus no diabetes (n = 15,673)

**e-Figure 6:**
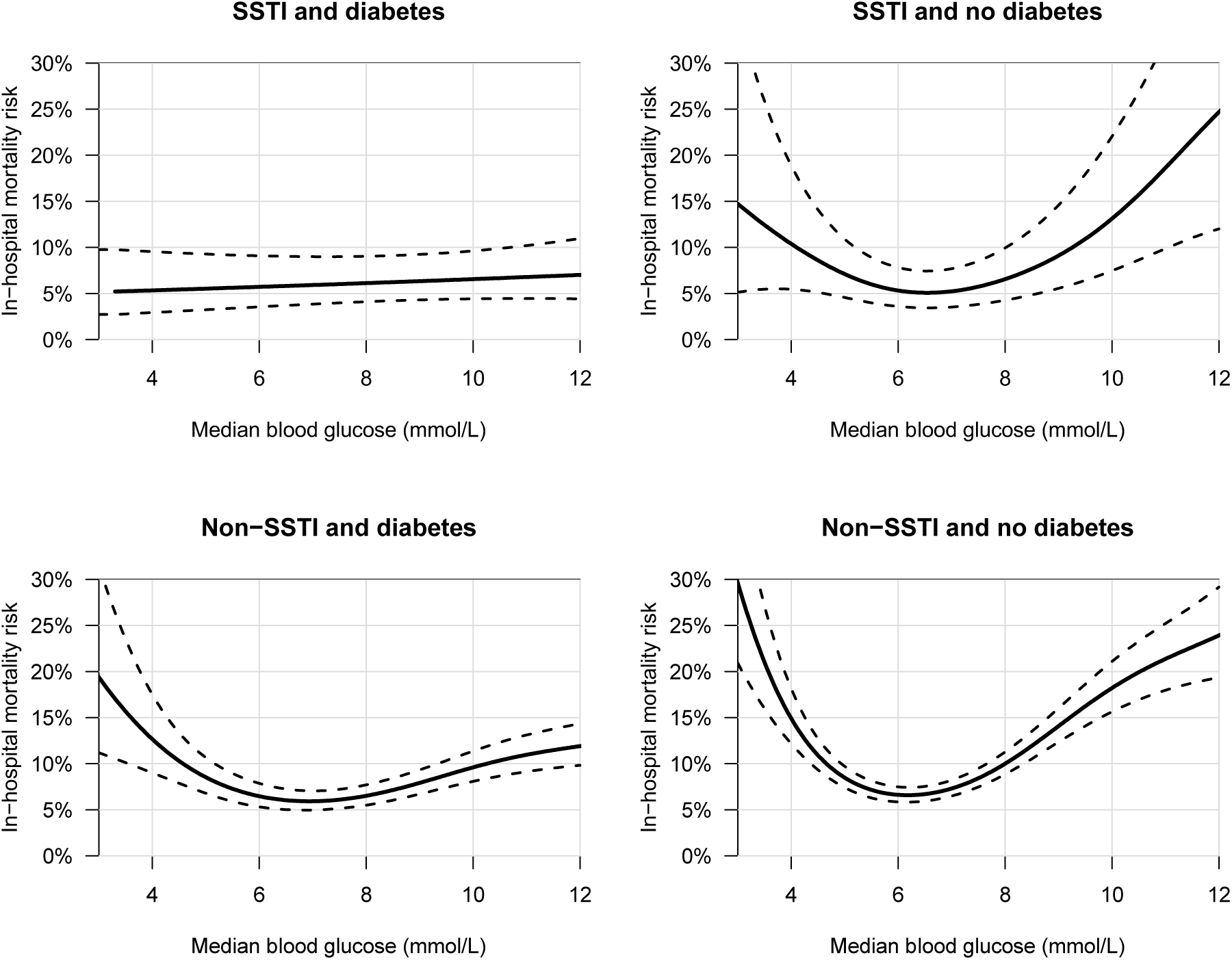
Subgroup analysis based on SSTI and diabetes. Diabetes and SSTI (*n* = 704), SSTI and no diabetes (*n* = 951), non-SSTI and diabetes (*n* = 5,997), non-SSTI and no diabetes (*n* = 14,722)

**e-Figure 7:**
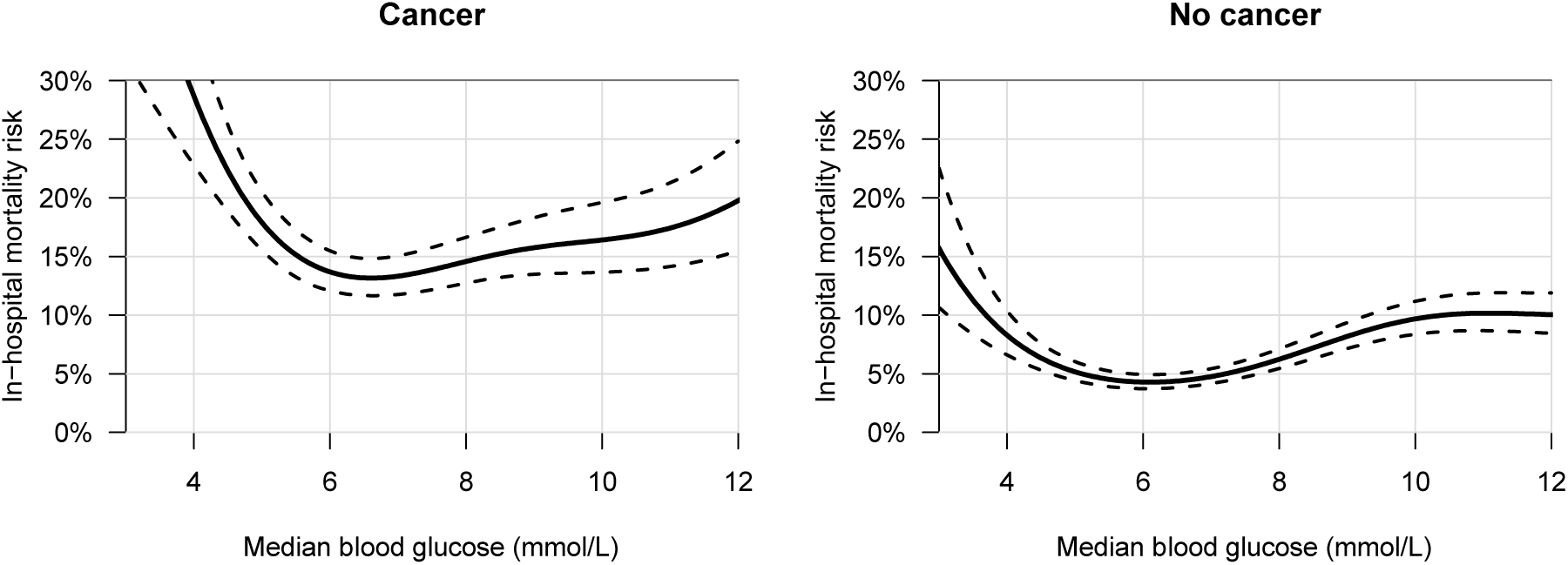
Subgroup analysis based on cancer. Cancer (*n* = 4,472) versus no cancer (*n* = 17,902)

**e-Figure 8:**
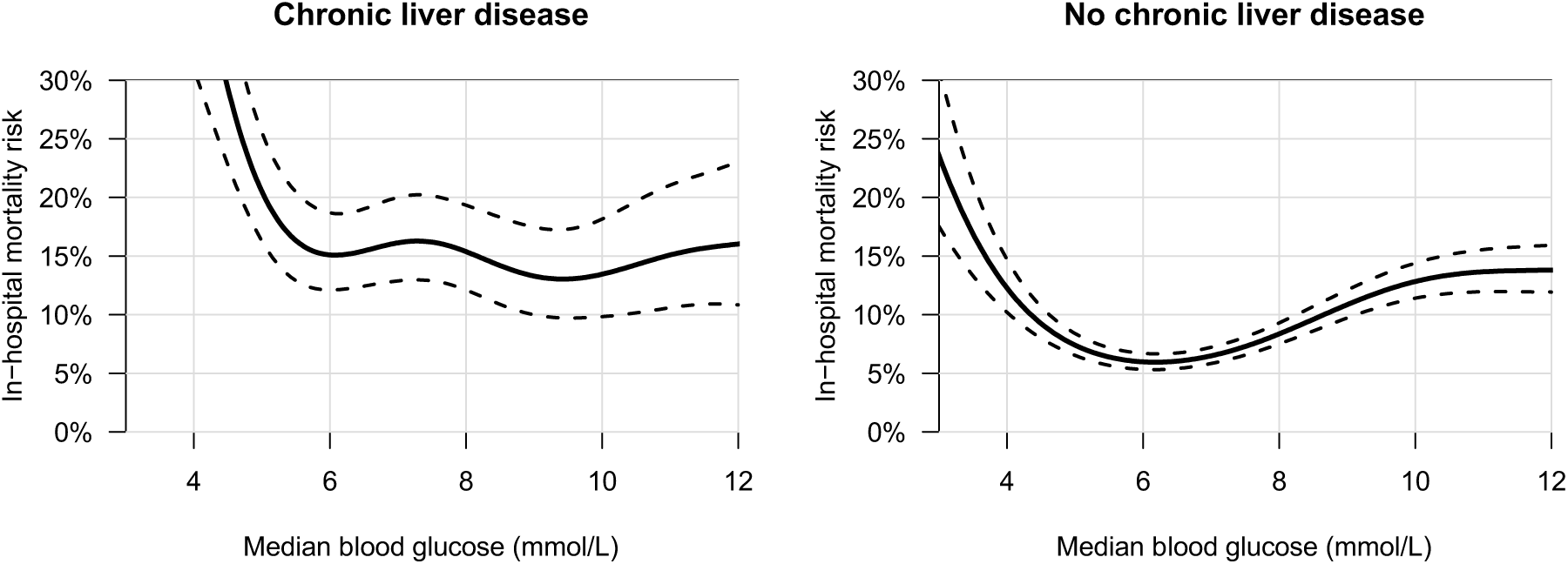
Subgroup analysis based on chronic liver disease. Chronic liver disease (*n* = 2,439) versus no chronic liver disease (*n* = 19,935)

**e-Figure 9:**
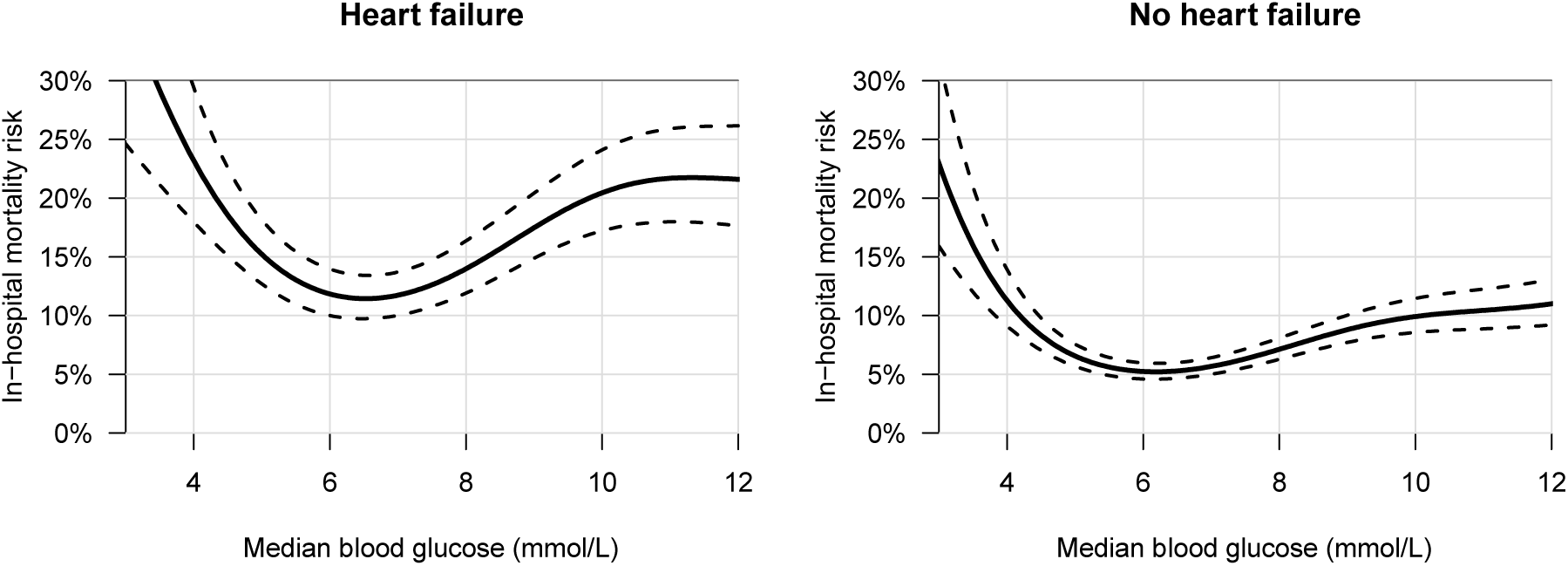
Subgroup analysis based on heart failure. Heart failure (*n* = 6,037) versus no heart failure (*n* = 16,337)

**e-Figure 10:**
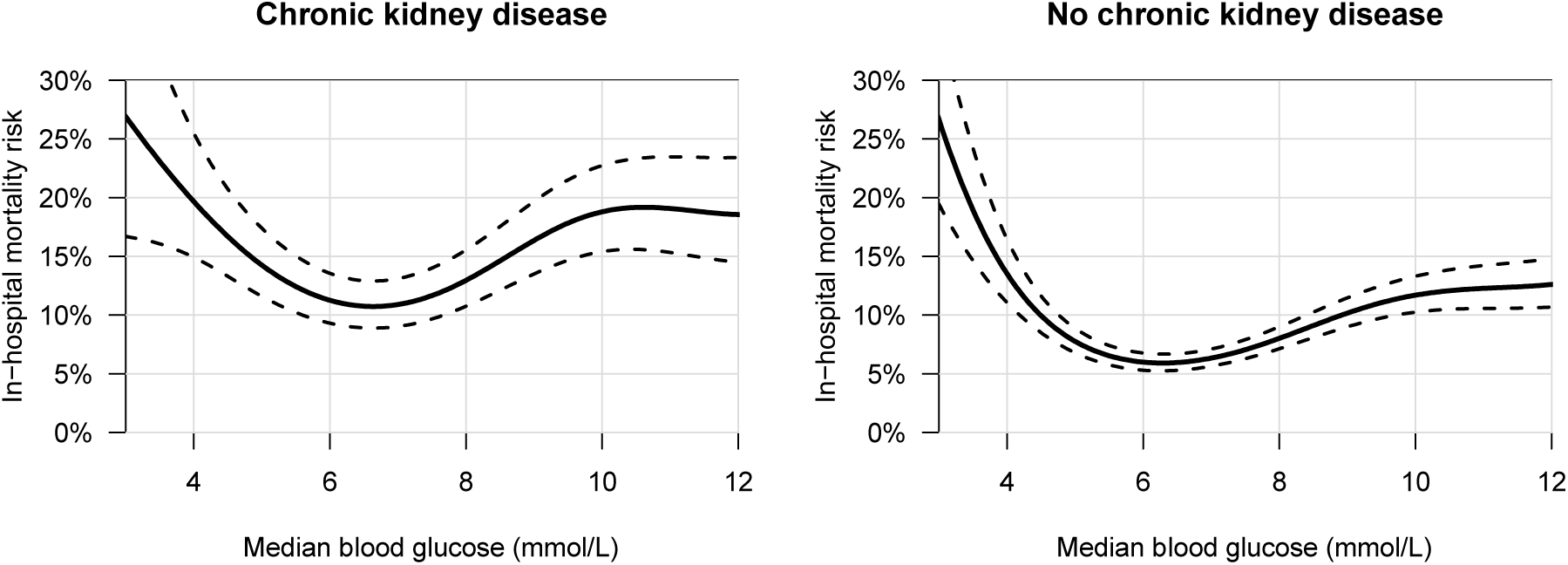
Subgroup analysis based on chronic kidney disease. Chronic kidney disease (*n* = 4,667) versus no chronic kidney disease (*n* = 17,707)

**e-Figure 11:**
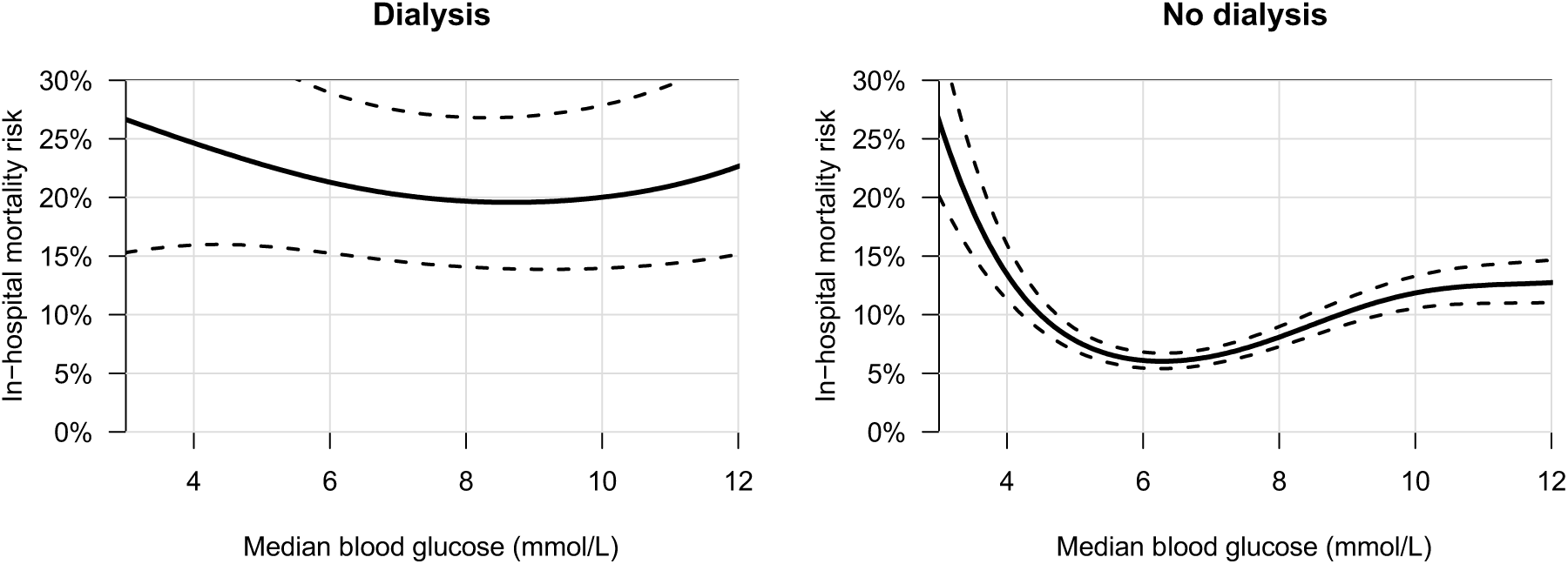
Subgroup analysis based on dialysis. Dialysis (*n* = 675) versus no dialysis (*n* = 21,699)

**e-Figure 12:**
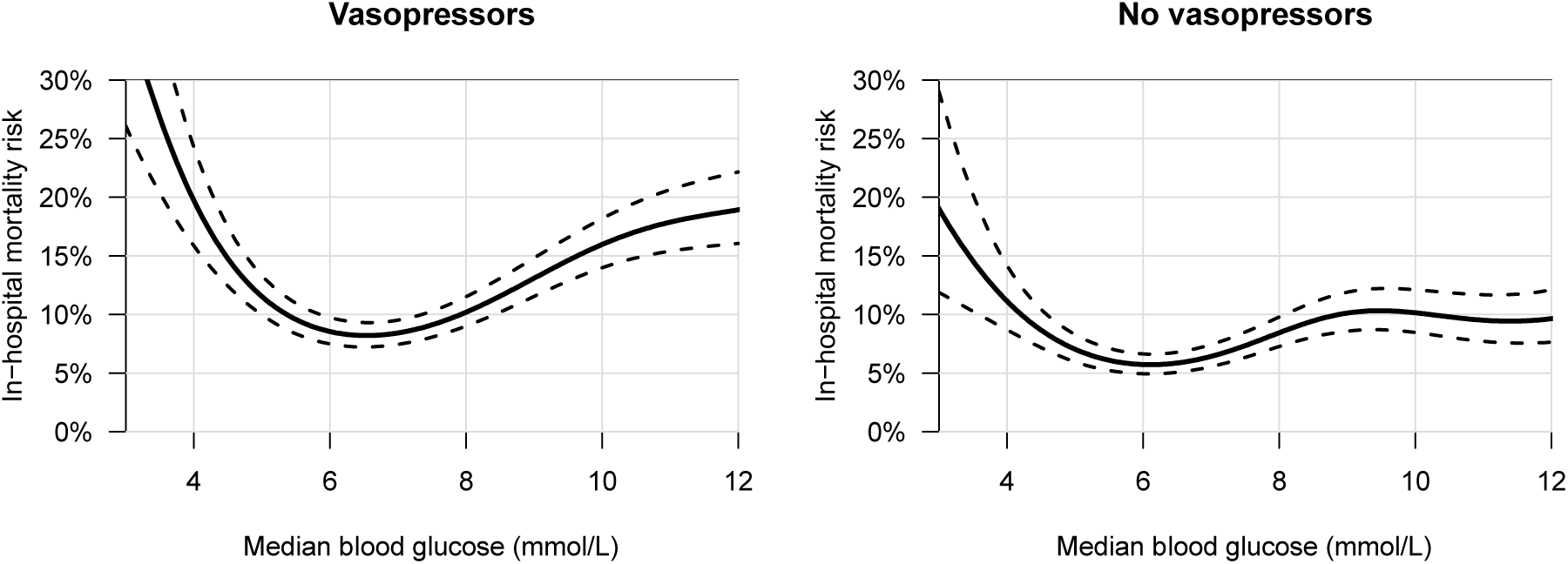
Subgroup analysis based on vasopressor use. Vasopressor use (*n* = 11,176) versus no vasopressor use (*n* = 11,198)

